# Beyond well-mixed: a simple probabilistic model of airborne disease transmission in indoor spaces

**DOI:** 10.1101/2021.12.05.21267319

**Authors:** Sijian Tan, Zhihang Zhang, Kevin Maki, Krzysztof J. Fidkowski, Jesse Capecelatro

## Abstract

We develop a simple model for assessing risk of airborne disease transmission that accounts for non-uniform mixing in indoor spaces and is compatible with existing epidemiological models. A database containing 174 high-resolution simulations of airflow in classrooms, lecture halls, and buses is generated and used to quantify the spatial distribution of expiratory droplet nuclei for a wide range of ventilation rates, exposure times, and room configurations. Imperfect mixing due to obstructions, buoyancy, and turbulent dispersion results in concentration fields with significant variance. The spatial non-uniformity is found to be accurately described by a shifted lognormal distribution. A well-mixed mass balance model is used to predict the mean, and the standard deviation is parameterized based on ventilation rate and room geometry. When employed in a dose-response function risk model, infection probability can be estimated considering spatial heterogeneity that contributes to both short- and long-range transmission.

## 1. Introduction

Understanding and predicting the transmission of infectious diseases is critical for assessing risk to individuals and informing policy changes. Airborne diseases, such as influenza, tuberculosis, measles, and SARS-CoV-2, most effectively spread in indoor settings, especially in spaces with poor ventilation and overcrowding. For example, it is now recognized that airborne transmission is the dominant transmission mode of SARS-CoV-2 during the COVID-19 pandemic (Zhang et al., 2020; Delikhoon et al., 2021), with the majority of super-spreading events occurring indoors (see, e.g., Zhang et al., 2020; Morawska et al., 2020; Shen et al., 2020; Miller et al., 2021).

The likelihood of infection depends on the exposure dose–the number of viral particles inhaled by a susceptible individual–which is a consequence of both short- and long-range transmission routes (Stilianakis and Drossinos, 2010; Liu et al., 2017; Wei and Li, 2016; Delikhoon et al., 2021). Short-range transmission is controlled by the initial advection of exhaled respiratory droplets, gravitational settling of larger droplets, and evaporation into droplet nuclei (Balachandar et al., 2020). Long-range transmission of droplet nuclei from a host to a susceptible person is governed by complex turbulent airflow, mechanical and passive ventilation, thermal buoyancy effects, and air filtration.

Due to their small size, expiratory droplet nuclei move passively with background air currents, and their concentration field, *C*(***x***, *t*), can be modeled by an advection-diffusion equation

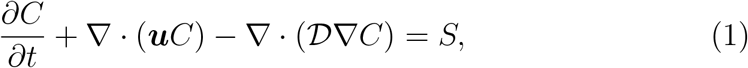

where ***u*** the background fluid velocity, 𝒟 is the diffusion coefficient, and *S* is a source term that accounts for pathogen inactivation and other contributions to changes in the concentration field. A direct solution to Eq. (1) is generally computationally expensive, particularly in the Navier–Stokes calculation of the background velocity. Thus, risk analyses that require a large number of iterations must rely on a simplified, or coarse-grained, representation.

Integrating Eq. (1) over the entire space results in an ordinary differential equation–the so-called well-mixed mass balance model–whose solution after an exposure time *t* with initially zero concentration is (Gammaitoni and Nucci, 1997)

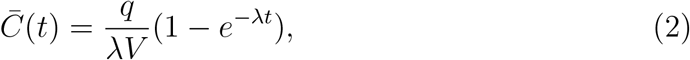

where *q* is the pathogen generation rate (by one or more infected sources) due to breathing, speaking, etc., *λ* is a loss coefficients that accounts for ventilation, deposition, etc., and *V* is the volume of the room. In this expression, 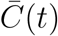 represents the average concentration field assuming a homogeneous environment in which expelled particles are uniformly distributed within the enclosed space. Under these assumptions, the total number of infectious particles inhaled by a susceptible individual (dose) is related to the concentration field according to 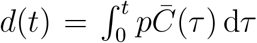 (Evans, 2020), where *p* is the pulmonary ventilation rate (breathing rate) of an individual.

When combined with an exponential dose-response function, the probability of infection, defined as the ratio of number of infections, *N*_*I*_, to number of susceptible individuals, *N*_*S*_, can be estimated according to

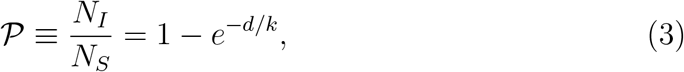

where *k* is a constant that depends on the infectiousness of the virus (Watanabe et al., 2010). Alternatively, the probability can be expressed in terms of the quantum of infection (or quanta) introduced by Wells (1955), which gives rise to the well-known Wells–Riley model (Riley et al., 1978). The aforementioned approaches act as the basis of most epidemiological models used today for assessing the risk of airborne diseases. For example, such models have been used to predict the risk of infection associated with measles (Riley et al., 1978), tuberculosis (Nicas, 1996; Gammaitoni and Nucci, 1997; Beggs et al., 2003), and more recently SARS-CoV-2 (Vuorinen et al., 2020; Morawska et al., 2020; Miller et al., 2021; Bazant and Bush, 2021).

A key drawback of the Wells–Riley model and dose-response functions based on a well-mixed mass balance is that the concentration of infectious material is assumed to be evenly distributed at each instant in time. It is well established that this is rarely true, even in spaces with high ventilation rates (Noakes and Sleigh, 2009; Anchordoqui and Chudnovsky, 2020; Zhang et al., 2021a). Consequently, infection risk due to short- and long-range exposure cannot be distinguished, and all susceptible individuals are treated as equally vulnerable. As pointed out in a review by Sze To and Chao (2010), *“[the] spatial distribution of airborne pathogens is one of the most important factors in infection risk assessment of respiratory disease*.*”*

A variety of approaches have been proposed to address the shortcomings of well-mixed models in recent years. Multi-zone (or zonal ventilation) models divide the space into sub-volumes (zones), each of which are assumed well-mixed with homogeneous composition (Axley, 1989; Bouia and Dali- cieux, 1991; Noakes and Sleigh, 2009). Susceptible individuals located in different zones are exposed to a different dose and thus experience different levels of infection risk. This approach is typically used in settings with multiple rooms or spaces with partitions, such as in hospitals (Nicas and Miller, 1999; Ko et al., 2001; Noakes et al., 2015), commercial airliners (Ko et al., 2004; Jones et al., 2009), and apartment buildings (Li et al., 2005). Several extensions to the multi-zone model have been proposed. Nicas (2000) developed a probabilistic model based on Markov chains, where each zone is treated as well-mixed but the concentration from one zone to another is described probabilistically, allowing for some variability in infection risk to be captured. Noakes and Sleigh (2009) proposed a stochastic version of the Wells–Riley model, where mixing between zones is limited. While multi-zone models have shown significant improvement over previous epidemiological models, the treatment of each zone as well-mixed precludes them from identifying infection to susceptible people in close proximity to the infectious source, unless detailed information regarding transport between interconnected spaces is known *a-priori* (Mora et al., 2003).

Sun and Zhai (2020) generalized the Wells–Riley model by introducing a distance index and a ventilation index to quantify the impact of social distance and ventilation effectiveness on the probability of infection. Guo et al. (2021) showed that this model yields reasonable accuracy against available data. However, they point out that it does not account for the location of infectors and fails to capture the spatial distribution of infection probability. Accordingly, Guo et al. (2021) introduced a so-called spatial flow impact factor into the Wells–Riley model and demonstrated success in predicting the spatial distribution of infection probability, which was used to identify optimal placement of individuals and facilities (e.g., air purifiers) in a simulated hospital ward. However, such an approach requires detailed flow measurements via experiments or computational analysis of the specific confined space prior to its use.

Computational fluid dynamics (CFD) has gained popularity in recent years for quantifying the spatial distribution of pathogens in confined spaces. Due to the wide range of length- and time-scales present in turbulent flows, obtaining the local velocity field, ***u***, in Eq. (1) via a direct solution to the Navier–Stokes equations is often not tractable except for early stage, short-range propagation of expiratory flows such as coughs or sneezes (e.g., Fabregat et al., 2021; Monroe et al., 2021; Chong et al., 2021). Instead, the mean flow field is often obtained via the Reynolds-averaged Navier-Stokes (RANS) equations, e.g., using a *k − ε* turbulence model (e.g., Noakes et al., 2006; Qian et al., 2009; He et al., 2011; Abuhegazy et al., 2020; Li et al., 2021; Zhang et al., 2021b). The concentration field obtained from CFD can then be employed in a dose-response function or used to inform multi-zone models (Gao et al., 2008; Qian et al., 2009; Zhang et al., 2021a). Due to its high computational cost, CFD is traditionally used to study specific scenarios under a limited number of permutations. Thus, its use for general risk assessment remains limited.

In this work, a general model is developed that accounts for spatial non-uniformity in indoor spaces. A large database of CFD results is generated for a wide range of representative spaces (see Fig. 1). The location of infected individuals, room geometry, ventilation rate, and exposure duration are varied to obtain a statistically significant representation of disease transmission in confined spaces. Details on the numerical simulations are provided in Sec. 2. A statistical analysis of the concentration field is then presented in Sec. 3, and a model is proposed for its probability density function. In Sec. 3.3, the model is employed in a dose-response function to predict the probability of infection via Monte Carlo sampling.

**Figure 1:**
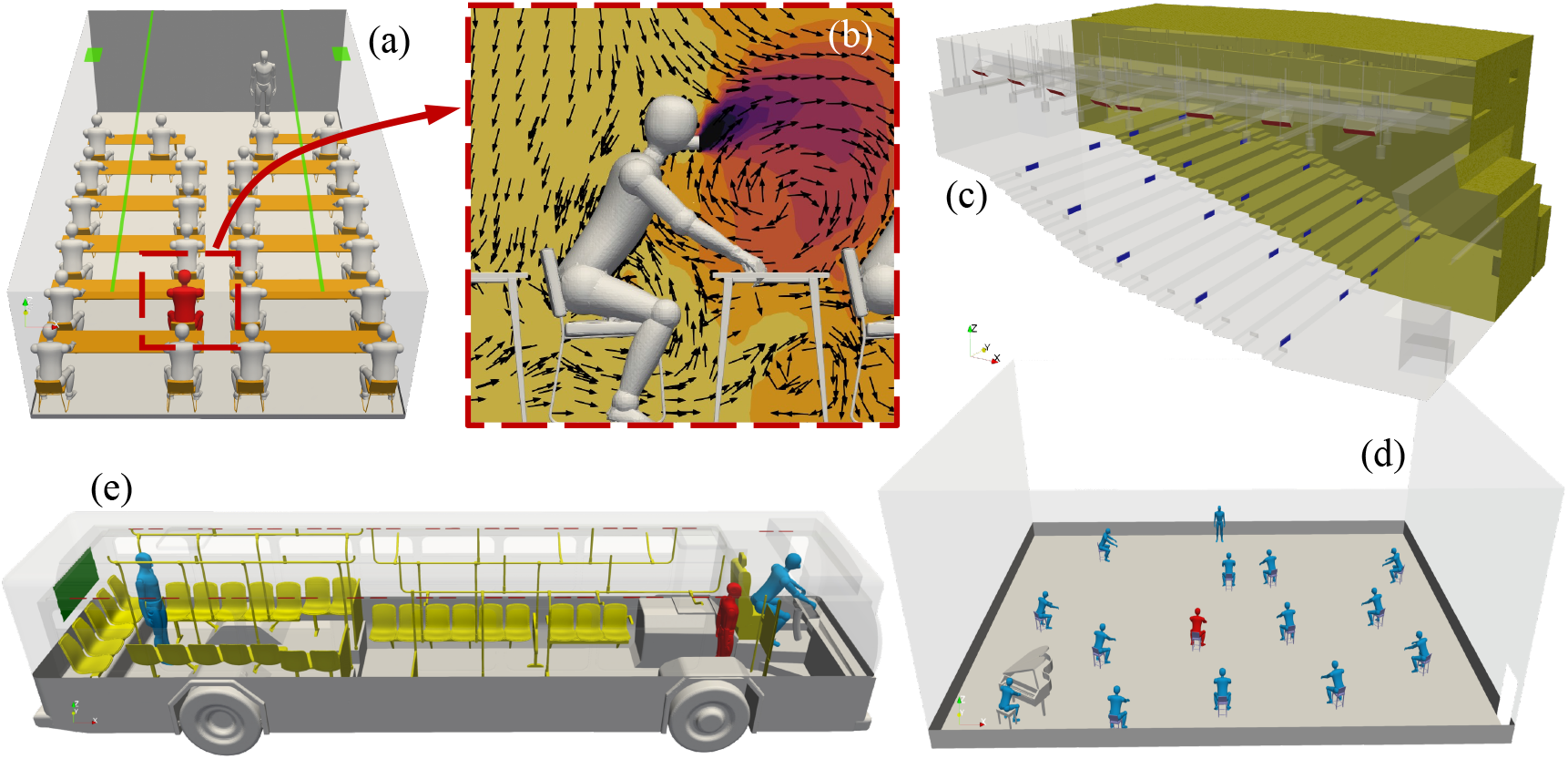
Schematic of the various geometries used in the numerical simulations. (a) Classroom. (b) Zoom-in of the infected individual (color shows concentration, arrows depict fluid streamlines). (c) Lecture hall. (d) Rehearsal hall. (e) Urban bus.

## 2. Simulation details

Simulations are conducted for several representative indoor spaces: an urban bus; a 30-student classroom; a large college-style lecture hall; and a music rehearsal space, as depicted in Fig. 1. Room dimensions and ventilation specifications are modeled after spaces at the University of Michigan. In each case, the computational domain is divided into a set of non-overlapping hexahderal cells. Local grid refinement is applied to small features, such as the return and supply vents, occupants, interior walls, tables, and seats. Details on the computational mesh and boundary conditions used in each case are summarized in Appendix A.

The airflow is assumed to be incompressible and turbulent, governed by

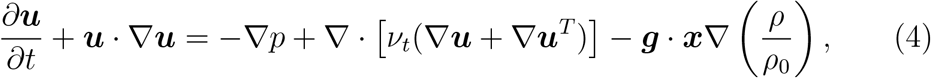

where *p* is the kinematic pressure, ***g*** is the acceleration due to gravity, and *ν*_*t*_ is the effective viscosity that accounts for both molecular and turbulent diffusion. The turbulent viscosity is obtained from the *k* − *ε* turbulence model (Launder and Spalding, 1983). Due to the small density variations of the air, the Boussinesq approximation is employed, so that density variations only appear in the gravitational term. The nominal air density is *ρ*_0_ = 1.2 kg/m^3^ and the local density, *ρ*, varies based on the temperature field according to the ideal gas law. The equation governing temperature is

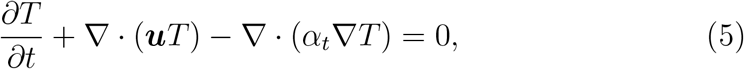

where *α*_*t*_ = *ν*_*t*_*/*Pr_*t*_ + *ν/*Pr is the effective thermal diffusivity, and Pr_*t*_ = 0.9 and Pr = 0.71 are the turbulent and laminar Prandtl numbers, respectively. The concentration field is solved according to Eq. (1), where the diffusion coefficient is given by 𝒟 = *ν*_*t*_*/*Sc_*t*_ + *ν/*Sc, where Sc_*t*_ = Sc = 1 are the turbulent and laminar Schmidt numbers, respectively.

For each case, a precursor simulation is performed to generate a fully-developed turbulent flow field inside the domain. The resulting flow field is used as the initial condition at which point the infected individual begins shedding aerosols. Occupants are represented by human-size manikins (standing and sitting) with uniform temperature of 32^*°*^C. The mouth and nose are modeled as an integrated round patch with a diameter of 4 cm and temperature of 34^*°*^C (see Fig. 1b). A turbulence intensity of 10% and a mixing length of 7.5 mm are enforced at the vicinity of mouth. In each simulation, it is assumed that only one infected person is present. Multiple simulations are run for each case, varying the location of the infector. A constant virus shedding rate of *q* = 50 s^*−*1^ is enforced as a boundary condition at the nose/mouth of the infected individual with a breathing rate of *p* = 6 l/min, corresponding to a highly contagious person speaking continuously and loudly (Vuorinen et al., 2020; Abkarian et al., 2020).

## 3. Results

In total, 174 simulations are performed: 163 of the 30-student classroom; eight of the lecture hall; one of the music hall; and two of the urban bus. Each indoor space is characterized by its volume *V*, height *H*, aspect ratio 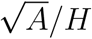, where *A* = *V/H* is the floor area, and ventilation rate denoted by the number of air changes per hour (ACH). The volume, ACH, and duration contribute to the mean concentration, 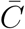, per Eq. (2), where the loss coefficient is related to ACH according to *λ* = ACH*/*3600. The variation in concentration field is found to depend on the room geometry 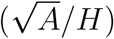 and relative position of the infector to the location of the return and supply vents (see Appendix B). Thus, for the analysis presented herein, averaging is performed over the volume of the indoor space Ω ∈ *V* and over different realizations by varying the location of the infector, according to

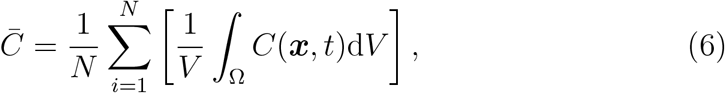

and

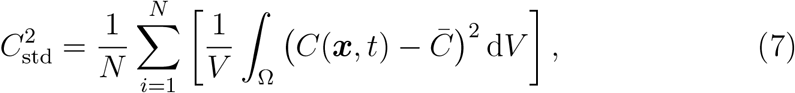

where 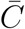 and *C*_std_ are the mean and standard deviation of the concentration field, respectively, *C*(***x***, *t*) is the local concentration at time *t*, and *N* are the number of realizations. It should be noted that in the region near the infector’s mouth, the concentration is extremely high and quickly drops before reaching susceptible individuals. Thus, computational cells containing the largest 0.1% values near the infector are removed.

### 3.1. Flow field and concentration distribution

The mean and standard deviation of the concentration field obtained from each case are summarized in Appendix B. Several important observations can immediately be made. First, the well-mixed mass balance model (2) yields an accurate prediction of the mean concentration when compared to the CFD results for all of the cases considered. Second, the variation about the mean, characterized by *C*_std_, is large, with values comparable to or larger than the mean. This has important consequences in risk assessment since the infection dose of an individual may vary significantly about the mean, depending on the relative proximity to the infector or supply and return vents. In addition, the variation in concentration is seen to be correlated with the room geometry: *C*_std_ tends to increase with increasing aspect ratio. Instantaneous snapshots of the airflow and concentration field in two of the classroom configurations are shown in Fig. 2. Turbulent mixing and recirculation are driven by the return and supply vents. The concentration field is diffused and convected by the air currents away from the infector to neighboring susceptible individuals. Because each case considers one infector, increasing the aspect ratio from 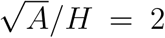 in Fig. 2(b) to 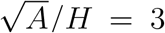 in Fig. 2(c) yields larger variation in the concentration field. This can be attributed to inadequate diffusion over the duration *t* droplet nuclei are being emitted. In the absence of any background airflow, transport of *C* is entirely controlled by diffusion. Because the diffusion time *A/ 𝒟 ≫t*, a larger surface area results in greater non-uniformity.

**Figure 2:**
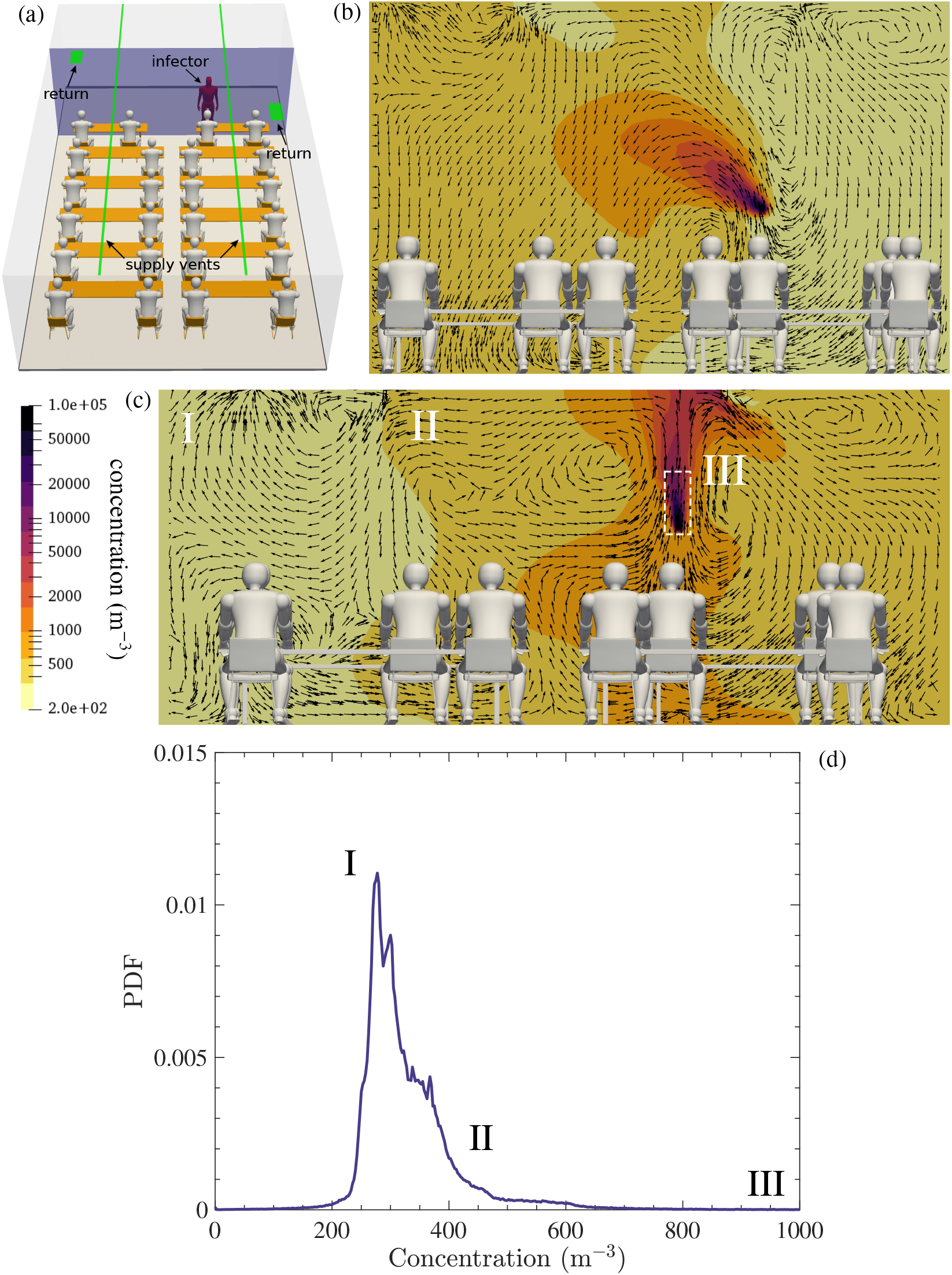
Concentration *C* contours and streamlines inside classrooms with ACH = 3 at different aspect ratios 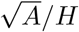. (a) Schematic of the classroom indicating location of the infector, supply vents, and return. (b) 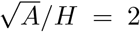. (c) 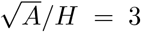. (d) PDF of concentration from (c) highlighting high probability events of moderate concentration (I), moderate probability of high concentration (II), and low probability of maximum concentration (III).

The probability density function (PDF) of concentration throughout the space in Fig. 2(d) shows (I) significant regions with concentration below the mean, corresponding to locations far from the infector; (II) high probability events of concentration near the mean value; and (III) a long tail with values significantly above the mean corresponding to regions near the infector. Thus, the PDF captures effects of both short-range (high *C*) and long-range (low and intermediate *C*) transmission.

Larger volume spaces, namely the lecture hall and music rehearsal hall considered here, exhibit vastly different behavior compared to the smaller classrooms. As shown in Fig. 3, the large height of the lecture hall results in significant vertical displacement of the concentration field. We attribute this behavior to buoyancy effects. The combined effect of convection by ventilation (or wind if windows are open) and buoyancy can be characterized by the densimetric Froude number according to 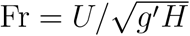 (Wykes et al., 2020), *where U* is a velocity scale associated with the supply vents (proportional to the ACH), and *g′* = *gβ*Δ*T*, where *β* = 1*/*293 K^*−*1^ is the thermal expansion coefficient of air at room temperature, and Δ*T* ≈ 14 K is the temperature difference between the infector and ambient air. If the ACH is sufficiently high to mix the space, the spatial variation in the concentration field will decrease. However, if there exists a significant temperature difference across the height of the room, the air can become stratified, trapping droplet nuclei near the ceiling (as shown in Fig. 3). For the cases considered herein, Fr ≪ 1. Thus, in spaces with large *H* (relative to the height of a person), the concentration distribution is expected to be controlled by buoyancy.

**Figure 3:**
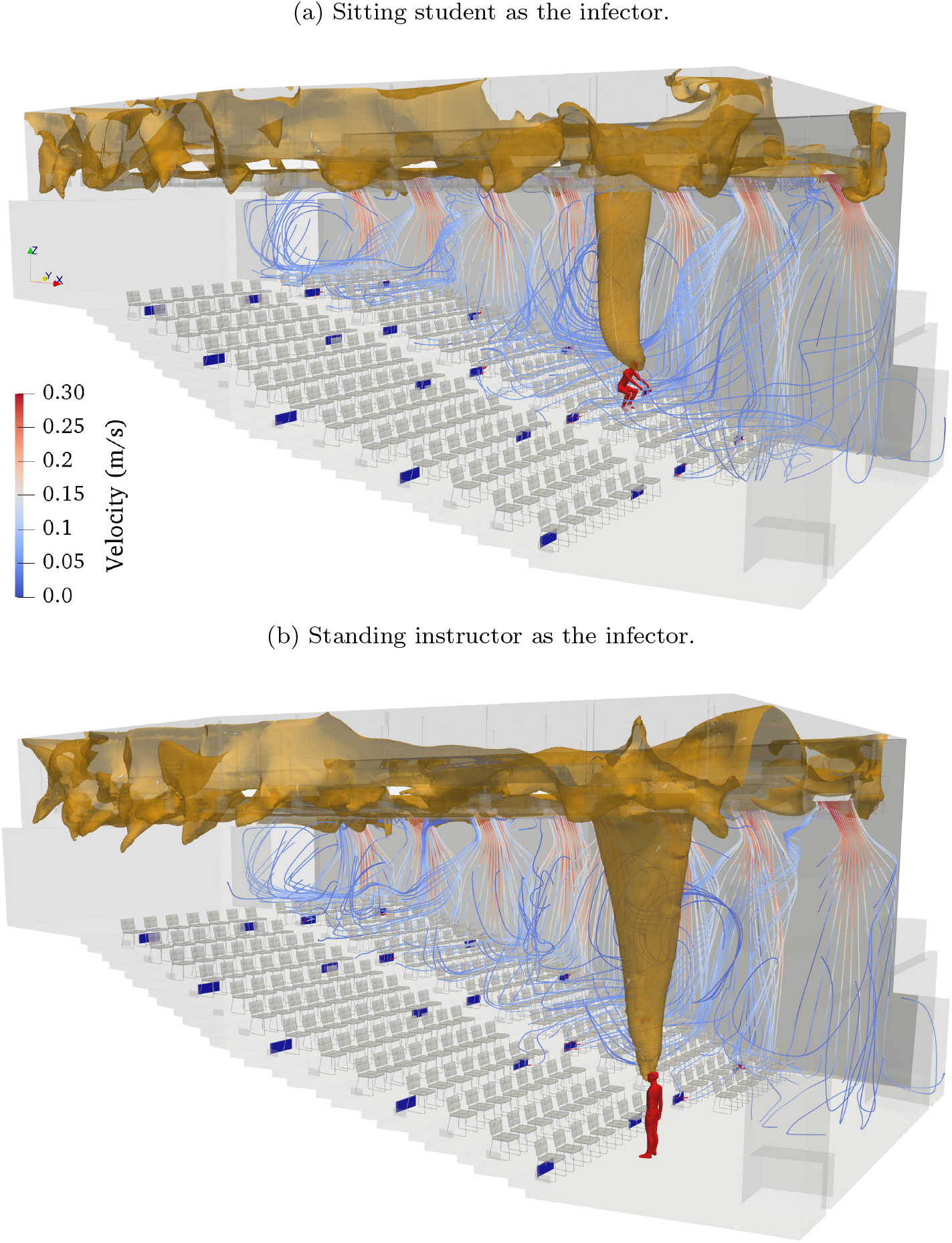
Instantaneous airflow streamlines and contour of concentration (*C* = 120 m^*−*3^) inside the lecture hall at *t* = 5400 s.

### 3.2. Probabilistic model

To capture the spatial non-uniformity in aerosol concentration for risk assessment, we propose to model the PDF based on a small number of input parameters that characterize the indoor space. A presumed-shape PDF is considered based on a shifted and scaled lognormal distribution. The two parameters that govern a standard lognormal distribution of a random variable *X* are *μ* and *σ*, corresponding to the mean and standard deviation of ln(*X*), respectively. The probability density function of *X* is

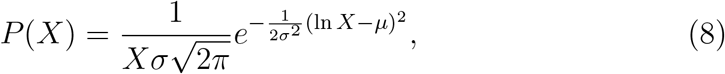

and the mean and standard deviation of *X* are given by 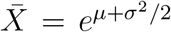 and 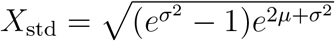, respectively. To model the concentration distribution, we scale and shift *X* to match the mean and standard deviation of the concentration field, *C*, according to

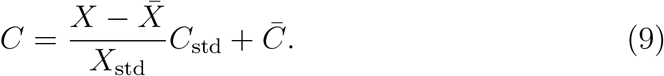

Solving for *X* in terms of *C* and substituting into Eq. (8) yields the probability density of *C*.

The parameters *μ* and *σ* govern the shape of the template lognormal distribution for the nondimensional variable, *X*. Based on fitting experiments with the CFD simulation dataset, we have found that *μ* = 0 and *σ* = 0.9 perform well in minimizing the error in the probability distributions. The PDF of *C* then only depends on the concentration mean, 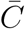, and the concentration standard deviation, *C*_std_. The well-mixed model per Eq. (2) is used for the former, and the latter is parameterized using the room geometry and ventilation parameters.

Linear regression of the CFD data of the classrooms, lecture hall, and rehearsal hall (see Table B.4) shows that a reasonable model for the standard deviation is given by

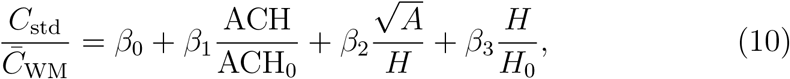

where 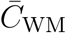 is the mean concentration obtained from the well-mixed mass balance (2), ACH_0_ = 1, *H*_0_ = 1 m, and the coefficients are *β*_0_ = −1.01, *β*_1_ = 0.11, *β*_2_ = 0.33, and *β*_3_ = 0.10. As described above, ACH captures the effects of turbulent mixing on the spatial distribution of the concentration field, while 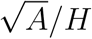 and *H* account for inadequate diffusion from the infector to the surroundings and buoyancy, respectively. We have found that a two-parameter model using ACH and 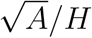 is sufficient for the classrooms, but inclusion of *H* is needed to capture the spatial non-uniformity in the lecture hall and music hall. The model given in Eq. (10) results in an *R*^2^ value of 0.97 over all of the cases listed in Table B.4. The model was developed for 0 ≤ ACH ≤ 6 h^*−*1^, 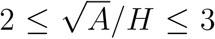, and 2.45 ≤ *H* ≤ 8.26 m, and therefore caution should be exercised when using values outside of this range.

As shown in Fig. 4, the normalized standard deviation increases linearly with aspect ratio. As noted earlier, ACH increases mixing in the room, resulting in smaller values of *C*_std_ in general. However, ACH also acts to dilute the air, resulting in a reduction of the mean concentration–recall ACH appears in the loss coefficient *λ* in Eq. (2). Consequently, *C*_std_*/C*_WM_ is observed to increase with ACH. Further comparisons between the standard deviation obtained from CFD with Eq. (10) are given in Table 1. Overall good agreement is observed over all of the indoor spaces considered. The largest discrepancy can be observed with the urban bus operated at ACH=16. As the regression was performed using ACH ∈ [0, 6], this value falls outside of the training set. It should also be noted that the buses were simulated using a clean air delivery rate of 20% (see Appendix A.4), and thus the ventilation rate used in the (10) is 5 times larger than the ACH used in predicting the mean.

**Table 1:** Predictions of *C*_std_ from CFD and the model given by Eq. (10).

| Description | ACH | $C_{\text{std}}$ (CFD) | $C_{\text{std}}$ (model) |
| --- | --- | --- | --- |
| Classroom | 1 | 335.26 | 329.56 |
| Classroom | 3 | 255.7 | 251.38 |
| Classroom | 6 | 233.56 | 211.58 |
| Lecture hall | 1 | 52.8 | 45.75 |
| Lecture hall | 6 | 20.24 | 19.44 |
| Rehearsal hall | 0 | 43.48 | 51.22 |
| Urban bus | 1.6 | 469.33 | 595.75 |
| Urban bus | 16 | 74.53 | 1903.6 |

**Figure 4:**
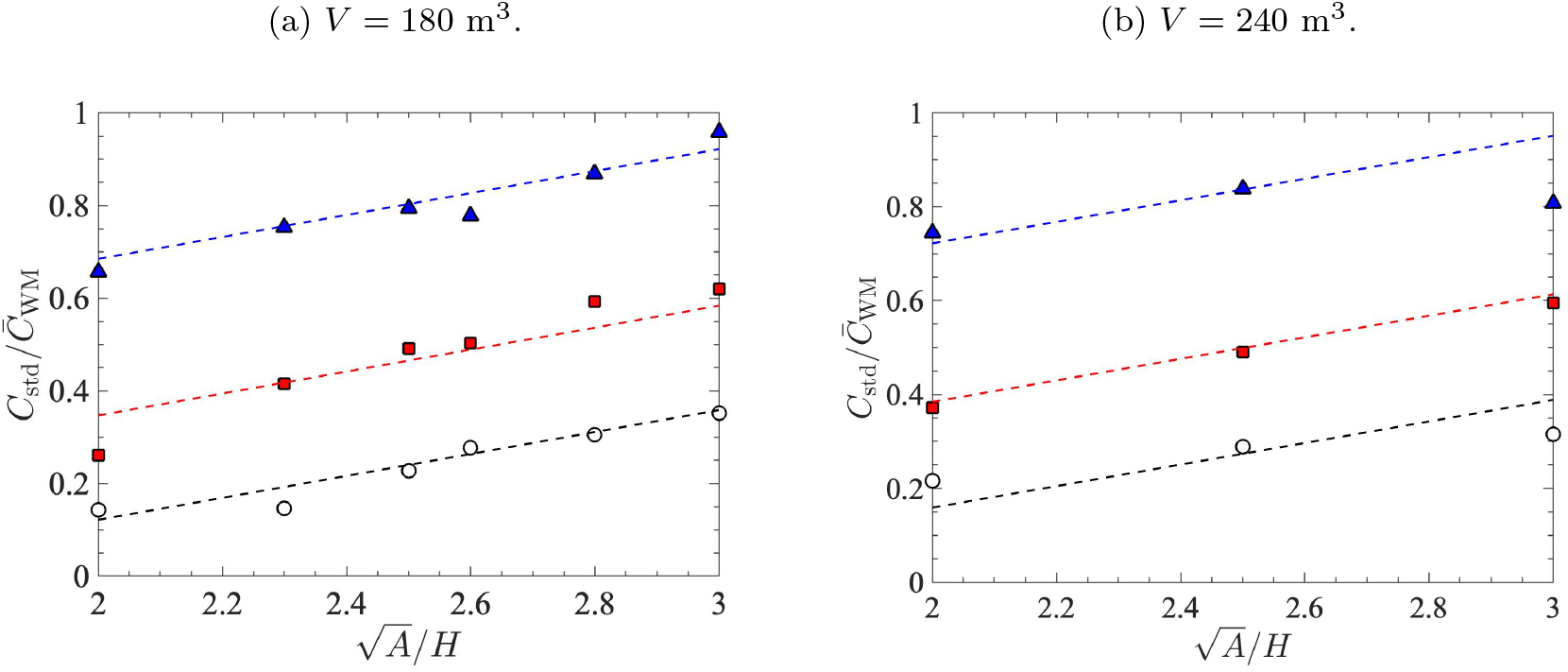
Standard deviation of the concentration field inside a classroom normalized by the mean obtained from a well-mixed mass balance. CFD results (symbols), model given by Eq. (10) (dashed lines) for ACH=1 (°), ACH=3 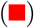, and ACH=6 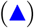.

Examples of the fully integrated PDF model, combining Eqs. (8)–(10) with *μ* = 0 and *σ* = 0.9, are shown in Fig. 5. For each case, the general trend observed in the CFD results is captured exceptionally well. The heavy-tailed nature of the distribution yields significant probability of concentration greater than the mean, corresponding to locations near the infector. Meanwhile, regions with local concentration below the mean are most probable, representative of long-range exposure. In the following section, the proposed model will be employed within a dose-response function to assess its impact on infection probability.

**Figure 5:**
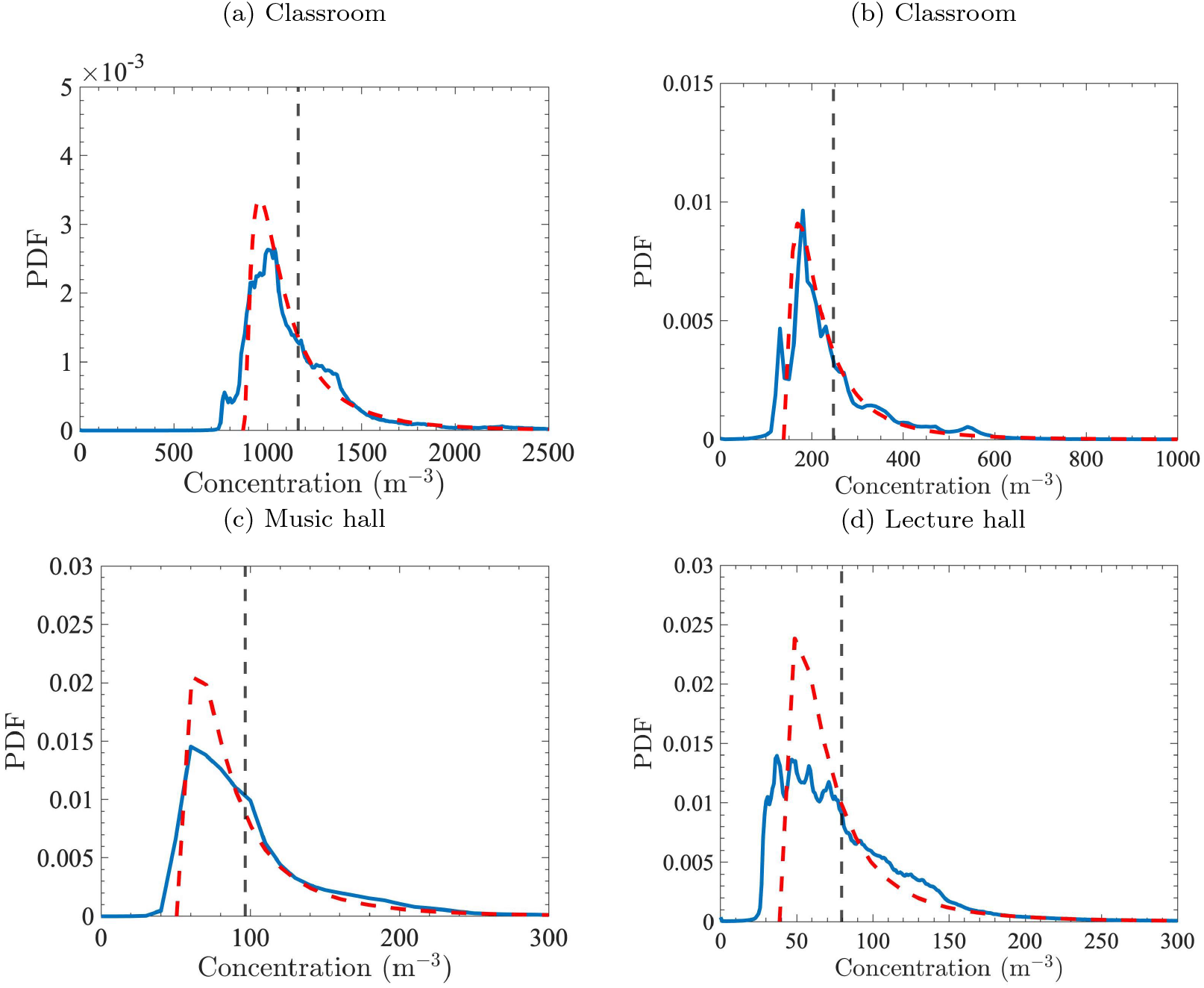
Concentration PDF obtained from CFD (***−***) and model using Eqs. (8)–(10) with *μ* = 0 and *σ* = 0.9 (***−−***). Mean obtained from the well-mixed mass balance (***−−***). Classroom with *V* = 120 m^3^ and ACH=1 averaged over *N* = 6 infector positions. (b) Classroom with *V* = 240 m^3^ and ACH=3, averaged over *N* = 3 infector positions. (c) Rehearsal hall with ACH=0 and *N* = 1. (d) Lecture hall with ACH=1, averaged over *N* = 4 infector positions.

### 3.3. Probability of infection

Dose response modeling is commonly used to estimate individual-level probability of infection based on the concentration of infectious aerosols (Watanabe et al., 2010; Sze To and Chao, 2010; Swanson et al., 2021). Here, we consider the exponential dose-response function given by Eq. (3), where the dose represents the total number of infectious particles an individual would inhale according to *d* = *pCt* (Evans, 2020). Here, the pulmonary ventilation rate is *p* = 1 × 10^*−*4^ m^3^/s (6 l/min), the concentration *C* is sampled from the proposed PDF model (9), and *t* is the duration. The parameter *k* typically varies between 75 and 500 (Watanabe et al., 2010; Swanson et al., 2021). Watanabe et al. (2010) found *k* = 410 provides a good fit for SARS coronavirus (SARS-CoV-1) based on data sets of infected mice, and this value is therefore used in the present study.

The shifted lognormal concentration PDF is sampled using a discrete inverse cumulative distribution function (CDF) method. *N* = 10^4^ random numbers *u*_*i*_ are generated from a uniform distribution between 0 and 1, and these are mapped to concentration values via *C*_*i*_ = CDF^*−*1^(*u*_*i*_), where the discrete CDF is evaluated by cumulative sums of the PDF values, and its inverse is computed by linear interpolation of point data.

Predictions of infection probability are shown in Fig. 6. In each case, a single infected person is presumed to reside within the space. The relative placement of the infector to susceptible individuals and to return and supply vents is accounted for in the modeled concentration PDF from which the samples are drawn. It can be seen that the probability of infection based on a dose obtained from the well-mixed mass balance is representative of the mean value obtained from sampling the concentration PDF. As expected, increasing ACH in the classroom leads to lower infection risk. Due to the large size of the music and lecture halls, infection probability is lower compared to the classroom despite having relatively low ACH.

**Figure 6:**
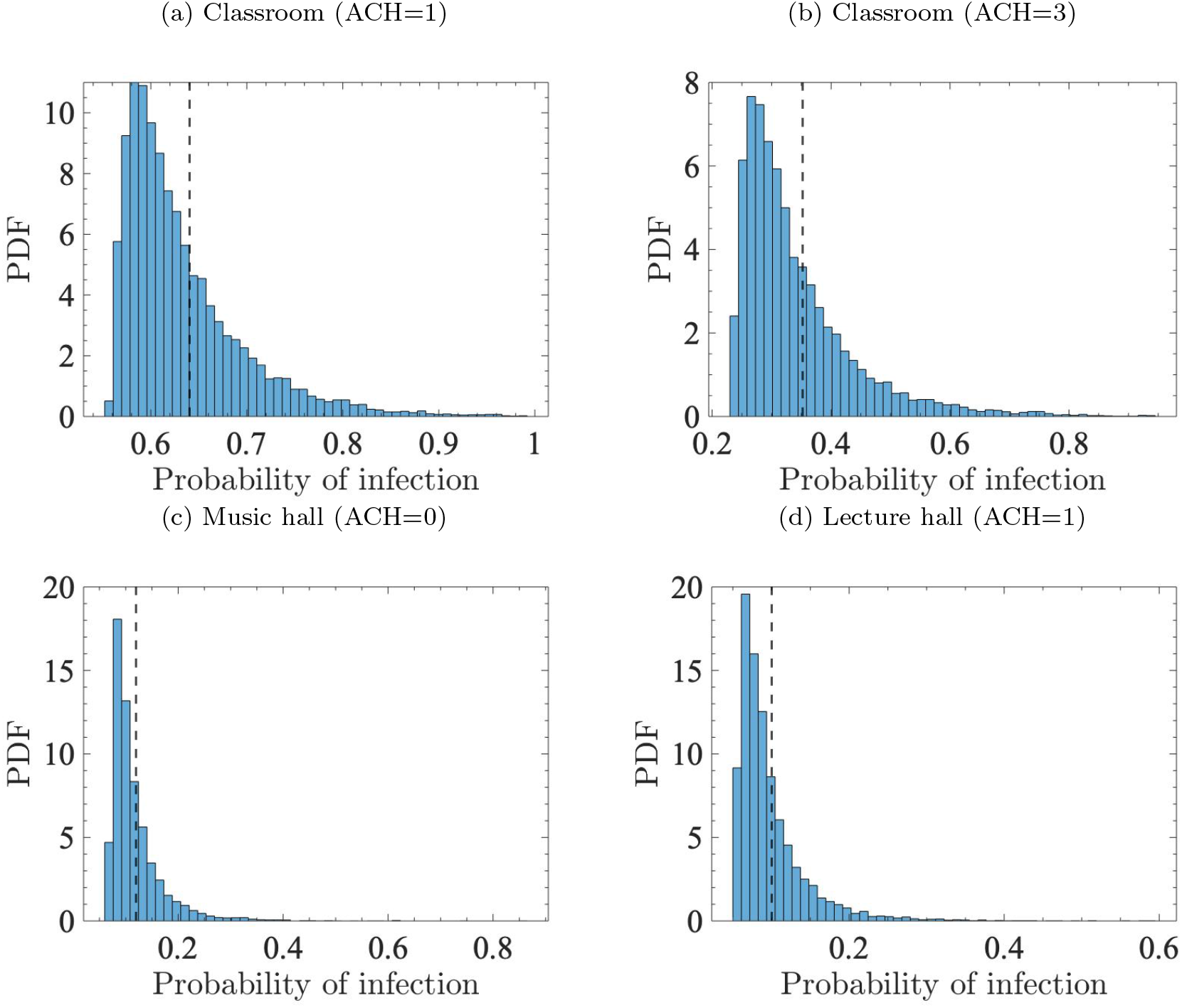
Histograms of the probability of infection in different scenarios. Each case consists of 10^4^ uniform random samples of the concentration PDF obtained from the proposed model. The vertical line denotes probability of infection using a dose obtained from the well-mixed mass balance. The classrooms have *V* = 180 m^3^ and 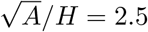.

Importantly, the probability of infection sampled from the proposed PDF model provides critical information that cannot be gleaned from a well-mixed assumption alone. Namely, the distribution of infection probability about the mean is large. A susceptible individual placed randomly within the confined space would experience high probability of having a lower infection risk. Yet, the same susceptible individual could be exposed to a much higher dose than predicted by a well-mixed mass balance. Therefore, upper and lower bounds of infection probability, or ideally the entire distribution, should be taken into account when determining safety guidelines for mitigating airborne transmission.

## 4. Conclusions

This work presents a simple probabilistic model that captures spatial heterogeneity of airborne pathogens. Dispersion of expiratory droplet nuclei in classrooms, lecture halls, and buses was simulated using the three-dimensional unsteady Reynolds-averaged Navier–Stokes equations. A database containing 174 simulations was generated and used to quantify the spatial distribution of droplet nuclei for a wide range of ventilation rates, exposure times, and room configurations.

The classical well-mixed mass balance was found to provide an accurate estimate of the mean concentration. However, the standard deviation in concentration was found to be relatively large, with values comparable to the mean. We attribute this spatial variation to three mechanisms: (i) turbulent mixing characterized by the ventilation rate; (ii) inadequate diffusion characterized by the lateral area of the room; and (iii) vertical displacement due to buoyancy in spaces with high ceilings.

The concentration distribution is modeled using a shifted and scaled lognormal distribution. Linear regression of the CFD data yields a simple expression for the standard deviation parameterized using the ventilation rate and room geometry. We demonstrate its use in an exponential dose-response function. By sampling the dose from the modeled concentration distribution, events corresponding to short-and long-range transmission are captured. This provides far more information on infection probability compared to a dose obtained from a well-mixed assumption.

Finally, we note the proposed model is complementary to many existing epidemiological models. For example, additional effects can be included in the well-mixed mass balance, such as deactivation due to ultraviolet lights or air filters (Evans, 2020). The concentration PDF can also be employed within multi-zone models to incorporate spatial heterogeneity within each zone. In addition, the effect of masks can be incorporated by adjusting the mean in the well-mixed mass balance and the breathing rate used in estimating the dose. Due to its low computational cost, the proposed model can be easily integrated into risk analyses for determining safety guidelines for mitigating airborne transmission, which to date have relied on well-mixed assumptions (e.g., Bazant and Bush, 2021; Swanson et al., 2021).

## Data Availability

All data produced in the present study are available upon reasonable request to the authors

## Acknowledgements

This work was supported by the College of Engineering at the University of Michigan. The computing resources and assistance provided by the staff of the Advanced Research Computing at the University of Michigan are greatly appreciated.

## Appendix A. Additional simulation details

Details on the numerical simulations performed in this study are provided below. A grid refinement study of the present configuration can be found in Zhang et al. (2021b).

**Table A.2:** Mesh resolution and the range of total number of cells.

| Scenario | Mesh resolution [mm] |  |  |  |  | # of cells<br>[million] |
| --- | --- | --- | --- | --- | --- | --- |
|  | Background | Supply vents | HVAC return | Mouth | Other surfaces |  |
| Classroom | 100 | 6.2 | 12.5 | 3.1 | 6.2 | 4.07-4.24 |
| Lecture hall | 100 | 12.5 | 12.5 | 3.1 | 25 | 6.47 |
| Rehearsal | 125 | — | — | 3.9 | 7.8 | 3.17 |
| Urban Bus | 125 | 2 | 31.2 | 3.9 | 7.8 | 5.86 |

**Table A.3:** Boundary conditions and material properties.

| Boundary name | Boundary conditions |
| --- | --- |
| HVAC supply vents | Velocity inlet with uniform flow, 25°C, 10% turbulence intensity and turbulence length scale is $5 \times 10^{-3}$ m |
| HVAC return | Pressure outlet |
| Mouth | Constant exhalation, 6 l/min, 34°C, 2.5% turbulence intensity, turbulence length scale is $7.5 \times 10^{-3}$ m and $\lambda = 50 \text{ s}^{-1}$ |
| Manikins | No-slip and 32°C |
| Other surfaces | No-slip and adiabatic walls |
| Material parameters | $\nu = 1.5 \times 10^{-5} \text{ m}^2/\text{s}$, $T_0 = 20^\circ\text{C}$, $\beta = 3 \times 10^{-3} \text{ K}^{-1}$, $\text{Pr} = 0.71$, $\text{Pr}_t = 0.9$, $g = 9.81 \text{ m/s}^2$, $\text{Sc} = 1$, $\text{Sc}_t = 1$. |

### Appendix A.1. Classroom

For the classroom simulations, three different volumes are considered: 120 m^3^, 180 m^3^ and 240 m^3^. For the 120 m^3^ cases, fixed dimensions (8.95 m × 5.46 m × 2.45 m) are used to analyze the effect of different positions of infectors and different ACH values. For the 180 m^3^ and 240 m^3^ cases, to analyze the effect of the geometry, multiple dimensions with different aspect ratio values are used (shown in Table B.4).

For all the classroom cases, similar HVAC systems are applied. The width of the inlet vents is 5 cm and the length of the inlet vents equals to the length of the classroom (shown in Fig 1(a)). The size of the outlet vents is the same for each case, which is 0.3 m × 0.6 m × 2 (two outlets). The maximum ACH value of this HVAC is 6 and different ACH values (1,3,6) are used for simulations.

### Appendix A.2. Lecture Hall

A stereolithography (STL) file is created using computer aided deisgn (CAD) software to represent the geometry of the Chrysler Lecture Hall at the University of Michigan (shown in Fig 1(c)), which has a large volume of 1753 m^3^. Manikins of 46 sitting students and a standing instructor, seats and instruments are placed inside the hall. Four different infector positions are chosen (one for each simulation). The entire simulation period is 90 minutes, where the sick person emits in the whole period. Different ACH values (1,6) are used for simulations.

### Appendix A.3. Music rehearsal hall

A rectangular box is adopted to represent the simplified geometry of a music rehearsal hall, which has a large volume of 2488 m^3^ with dimensions of 19.82 m × 15.24 m × 8.24 m (*L* × *W* × *H*). Manikins of 14 sitting students and a standing conductor, seats and instruments are placed inside the hall. The entire simulation period consists of two 40-min classes with a 20-min break in between, where the sick person only emits during classes. No HVAC system is present in this hall, the airflow is mainly driven by pulmonary ventilation of the occupants and temperature gradients. A door is set to balance the pressure inside the domain.

### Appendix A.4. Urban bus

For the bus simulations, an urban bus that is used on the campus of the University of Michigan is studied. The geometry, including the interior of the cabin, windows, doors, seats, handrails, ventilation supply and return are determined from a laser scanner and used for generating the computational grid of the fluid domain. A rendering of the bus is shown in Fig 1 (e). The bus dimensions are 12.1 m × 2.58 m × 2.95 m (*L* × *W* × *H*) with a total interior volume of 52 m^3^. A total of 42 supply vents are located along both sides of the bus ceiling and has an orientation such that air exits vertically downward. The single ventilation fan draws air from the passenger compartment through a return vent in the back of the bus, and adds 20% fresh air from outside before returning the air to the cabin through supply vents. Each supply vent has a dimension of 9 in by 1 in (0.229 m by 0.0254 m), and the single return vent is 4 ft by 1.5 ft (1.22 m by 0.457 m). This HVAC system can provide a maximum flow rate of 2,500 ft^3^ /min (70.8 m^3^ /min), which is equivalent to an ACH of 16 considering the fresh air rate of 20%. With such ventilation system, the airflow moves up and down in transverse direction and the net flow is rearward through the compartment. Manikins are placed at different locations inside the bus: a driver sitting behind the wheel and standing passengers. Further details can be found in (Zhang et al., 2021b).

## Appendix B. Simulation results

**Table B.4:**
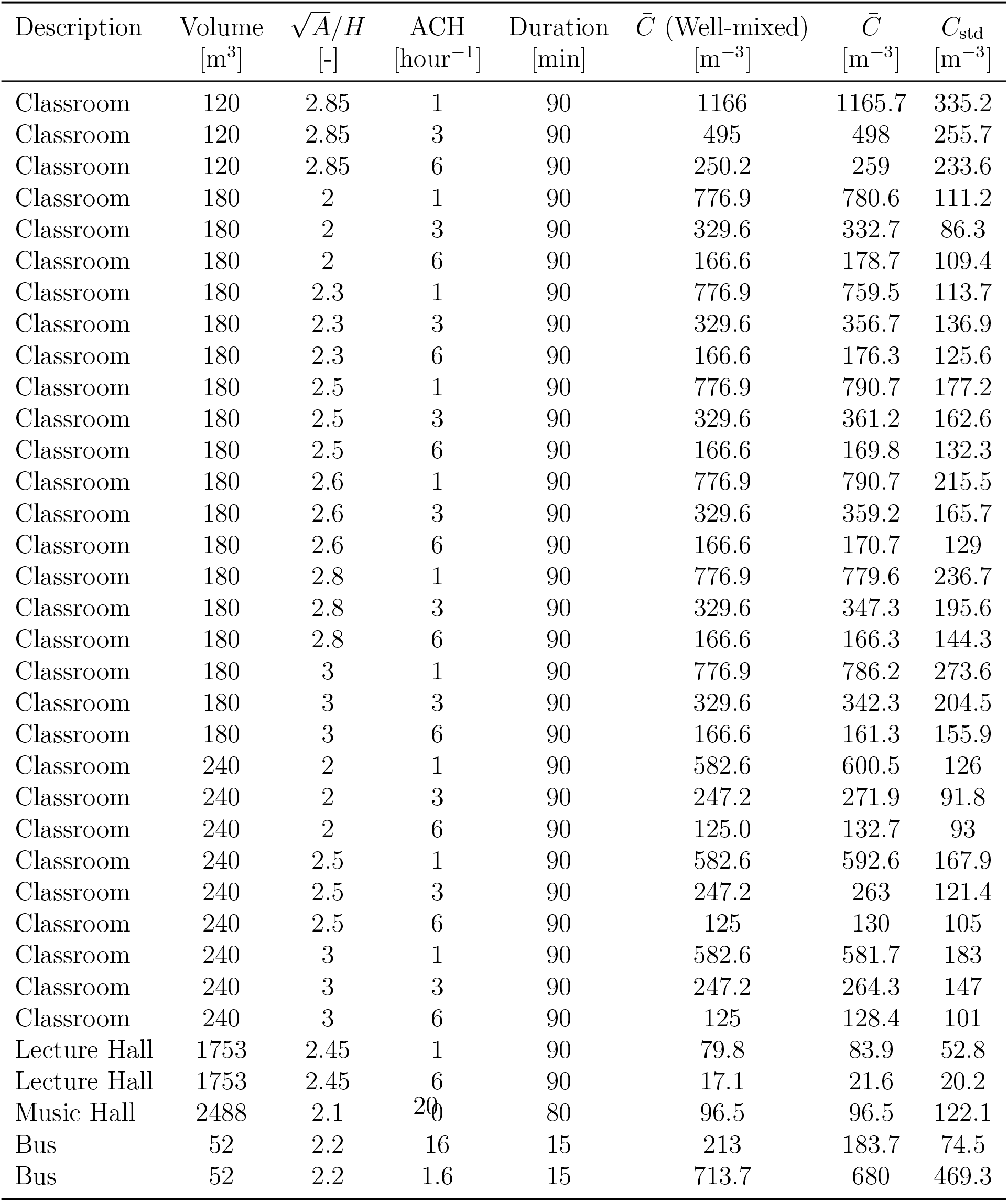
Mean and standard deviation of the concentration field obtained from CFD. Mean obtained from the well-mixed mass balance (2) shown for reference.

